# Real-world treatment patterns and clinical outcomes in Korean patients with AML ineligible for first-line intensive chemotherapy: A subanalysis of the CURRENT study, a non-interventional, retrospective chart review

**DOI:** 10.1101/2022.06.15.22276442

**Authors:** Soo-Mee Bang, Ka-Won Kang, Ik-Chan Song, Alexander Delgado, Cynthia Llamas, Yinghui Duan, Ji-Young Jeong, Je-Hwan Lee

## Abstract

**Background:** Although most elderly patients with acute myeloid leukemia are ineligible for intensive chemotherapy, treatment options remain limited. CURRENT (UMIN000037786), a real-world, non-interventional, retrospective chart review, evaluated clinical outcomes, clinicopathologic characteristics, and treatment patterns in these patients. We present results from a subanalysis of Korean patients in this study.

**Methods:** Patients were aged ≥18 years with primary or secondary acute myeloid leukemia ineligible for intensive chemotherapy who initiated first-line systemic therapy or best supportive care between 2015 and 2018 across four centers in Korea. Primary endpoint was overall survival from diagnosis. Secondary endpoints included progression-free survival, time to treatment failure, and response rates. Data analyses were primarily descriptive, with time-to-event outcomes estimated using the Kaplan-Meier method, and Cox regression used to determine prognostic factors for survival.

**Results:** Among 194 patients enrolled, 84.0% received systemic therapy and 16.0% received best supportive care. Median age at diagnosis was 74 and 78 years, and Eastern Cooperative Oncology Group performance status 0 or 1 was reported in 73.0% and 48.4% of patients, respectively; poor cytogenetic risk was reported in 30.1% and 16.1% of patients. Median overall survival was 7.83 versus 4.50 months, and median progression-free survival was 6.73 versus 4.50 months in the systemic therapy versus best supportive care groups. Prognostic factors (all *P* <0.05) affecting overall survival included secondary acute myeloid leukemia (hazard ratio, 1.67 [95% CI: 1.13–2.45]), Eastern Cooperative Oncology Group performance status ≥2 (2.41 [1.51–3.83]), poor cytogenetic risk (2.10 [1.36–3.24]), and Charlson comorbidity index ≥1 (2.26 [1.43–3.58]).

**Conclusion:** Clinical outcomes are poor in Korean patients with acute myeloid leukemia ineligible for intensive chemotherapy who are prescribed current systemic therapies or best supportive care. There is a substantial unmet need for novel agents (monotherapy or in combination) to improve clinical outcomes in this patient population.

## Introduction

Acute myeloid leukemia (AML) is a hematologic malignancy characterized by the rapid proliferation of abnormally differentiated myeloid blast cells [1]. AML, the most common type of leukemia in adults worldwide [2], predominantly affects elderly individuals, with about 60% of patients diagnosed at ≥65 years of age [3]. From 1990 to 2017, the global incidence of AML rose by 87%, with 119,570 cases recorded in 2017 [4]. In Korea, AML is the most frequently diagnosed myeloid malignancy and is most prevalent in patients aged 60 to 79 years [5]. Despite the greater prevalence of AML in older versus younger adults, survival outcomes for this population remain extremely poor [6].

The current standard of care for AML is intensive chemotherapy (ICT), but approximately 50% of patients are ineligible for this treatment [7] owing to factors such as advanced age, poor performance status, and prevalence of comorbidities [8, 9]. AML-related genetic abnormalities can also increase the likelihood of resistance to ICT [9]. Treatment options for these patients remain limited and include low-intensity treatment with hypomethylating agents (HMAs), low-dose cytarabine (LDAC), and best supportive care (BSC) [2, 10]. The availability of targeted therapies, such as inhibitors of B-cell lymphoma–2 (BCL-2), isocitrate dehydrogenase isoforms 1/2 (IDH1/2), FMS-like tyrosine kinase–3 (FLT3), and Hedgehog (Hh), is also increasing for patients who are ineligible for ICT [11].

Prognostic models have been developed to determine the suitability of older patients for ICT, yet there is no consensus regarding their optimal treatment [12-14]. Treatment decision-making for elderly patients with AML is an escalating global clinical challenge in light of emerging new agents and is compounded by an increasing incidence of AML due to the aging population [4, 15]. Thus, there is a growing need to understand current treatment strategies and their associated clinical outcomes in patients who are ineligible for ICT.

The CURRENT study was an international, real-world, non-interventional, retrospective chart review that aimed to evaluate clinical outcomes, clinicopathologic characteristics, and treatment patterns of patients with AML deemed ineligible for ICT [16]. Here, we report that clinical outcomes were poor among the subgroup of Korean patients included in the CURRENT study.

## Methods

### Study design

The CURRENT study [16] enrolled 1792 patients across 112 community or hospital medical centers from 22 countries between January 1, 2015, and December 31, 2018; four of the medical centers were in Korea. Notification was made to the responsible ethics committees, health institutions, and/or competent authorities as required by local laws and regulations.

Ethics committee approval was obtained for this study, with the following institutional review board approval numbers (Seoul National University Bundang Hospital: B-1908/559-102; Korea University College of Medicine: K2019-1535-001; Chungnan National University School of Medicine: 2019-09-027; Asan Medical Center: S2019-1692-0001). Data collection was carried out anonymously, and final data cut-off was March 31, 2020.

### Study population

Eligible patients were aged ≥18 years, diagnosed with primary or secondary AML, and ineligible for ICT based on physician assessment of age, Eastern Cooperative Oncology Group (ECOG) performance status, comorbidities, regional guidelines, and institutional practice. Patients were also required to have commenced first-line systemic therapy with low-intensity chemotherapy (e.g. HMAs, including azacytidine and decitabine, or LDAC), targeted therapy, or BSC and to have attended at least two practice visits to the physician during the treatment period in addition to the initial treatment visit. Exclusion criteria included undiagnosed AML, acute promyelocytic leukemia, and having received first-line therapy for AML in a clinical trial. Patients were followed up until the last recorded contact or death (whichever came first), and all visits were completed before data extraction.

### Endpoints

The primary endpoint was overall survival (OS; measured from diagnosis of AML). Secondary endpoints included progression-free survival (PFS), time to treatment failure (TTF), response rate (including complete remission [CR] and CR with incomplete hematologic recovery [CRi]), and duration of response (DoR).

### Data collection

Anonymized patient data including age, sex, disease characteristics, prior treatment, ECOG performance status, cytogenetic risk, and Charlson comorbidity index (CCI) were extracted from patient charts and/or site documentation, and recorded via electronic case report forms (CRFs) completed by each center.

### Sample size

Target sample size for the overall CURRENT study was 1600 patients, and the target sample size in Korea was 170 patients. Because of the descriptive nature of the study, formal statistical power considerations are not provided. However, the sample size was considered sufficient to provide reasonably precise estimates.

### Statistical analyses

Data analyses were primarily descriptive. Continuous variables were described using mean, standard deviation (SD), median, and ranges. Categorical variables were reported as counts and proportions. Time-to-event data were estimated using the Kaplan-Meier method, with median time and 95% confidence intervals (CIs) reported. Log-rank test or Wilcoxon test were used to compare Kaplan-Meier estimates of survival between patient subgroups. Cox regression analyses were performed to evaluate the association between patient variables and estimates of median OS and PFS. Missing data were captured via an “unknown” option in the electronic CRFs wherever appropriate. No imputation was performed, and all analyses were conducted on available data only.

## Results

### Patient demographics and clinical characteristics

At final data cut-off, 194 Korean patients were enrolled. Patient baseline characteristics by treatment group are provided in Table 1. In the first-line systemic therapy and BSC groups, respectively, median age was 74.0 and 78.0 years, 64.4% and 48.4% of patients were male, and secondary AML was diagnosed in 25.2% and 29.0% of patients. The majority (73.0%) of patients in the first-line systemic therapy group had an ECOG performance status of 0 or 1; in the BSC group, approximately half (51.6%) had an ECOG performance status ≥2.

**Table 1.** Baseline demographics and patient characteristics.

|  | First-line systemic therapy |  |  | BSC<br>(n = 31) | *P* value |
| --- | --- | --- | --- | --- | --- |
|  | All<br>(n = 163) | HMA<br>(n = 145) | LDAC &<br>other<br>(n = 18) |  |  |
| **Male** | 105 (64.42) | 89 (61.38) | 16 (88.89) | 15 (48.39) | 0.0186* |
| **Age at diagnosis,**<br>**median (range),**<br>**years** | 74 (53–87) | 74 (53–87) | 72 (61–82) | 78 (46–87) | 0.0500† |
| >75 | 61 (37.42) | 56 (38.62) | 5 (27.78) | 20 (64.52) | 0.0133* |
| **Secondary AML** | 41 (25.15) | 35 (24.14) | 6 (33.33) | 9 (29.03) | 0.2654‡ |
| MDS | 22 (53.66) | 17 (48.57) | 5 (83.33) | 7 (77.78) | 0.6057‡ |
| CMML | 7 (17.07) | 6 (17.14) | 1 (16.67) | 1 (11.11) | – |
| MPN | 7 (17.07) | 7 (20.00) | 0 | 0 | – |
| t-AML | 5 (12.20) | 5 (14.29) | 0 | 1 (11.11) | – |
| **Prior HMA Tx for**<br>**antecedent disorder** | 8 (19.51) | 3 (8.57) | 5 (83.33) | 6 (66.67) | <0.0001‡ |
| **ECOG**<br>**performance status** |  |  |  |  |  |
| 0–1 | 119 (73.01) | 109 (75.17) | 10 (55.56) | 15 (48.39) | 0.0059* |
| ≥2 | 44 (26.99) | 36 (24.83) | 8 (44.44) | 16 (51.61) | – |

Data are n (%) unless otherwise stated.
*P* value indicates statistical difference in a three-way comparison between BSC, HMA, and LDAC and other systemic therapies.
AML, acute myeloid leukemia; BSC, best supportive care; CMML, chronic myelomonocytic leukemia; ECOG, Eastern Cooperative Oncology Group; HMA, hypomethylating agent; LDAC, low-dose cytarabine; MDS, myelodysplastic syndrome; MPN, myeloproliferative neoplasm; t-AML, therapy-related AML; Tx, treatment.
\*Chi-squared test; †Kruskal-Wallis test; ‡Fisher's exact test.

Cardiovascular, pulmonary, liver, renal, and other comorbidities were reported in 130 (89.7%), 18 (100.0%), and 28 (90.3%) patients who received HMA, LDAC and other systemic therapies, and BSC, respectively (S1 Table).

Patient molecular profiling and cytogenetic risk data by treatment groups are provided in S2 Table. Of the patients who received first-line systemic therapy with available cytogenic risk data (n = 145), 66 (45.5%), 30 (20.7%), and 49 (33.8%) had favorable, intermediate, and poor risk, respectively, according to the cytogenetic risk classification in the CRF (S3 Table). Of 16 patients who received BSC with available cytogenic risk data, the respective risk proportions were seven (43.8%), four (25.0%), and five (31.2%) patients. Of the patients who received first-line systemic therapy with available molecular data (obtained using next-generation sequencing or targeted mutation testing; n = 144), 49 (34.0%) had a mutation. None of the patients who received BSC with available molecular data (n = 22) had mutations.

Patients who received first-line systemic therapy were more likely to be <75 years of age compared with the BSC group (62.6% vs 35.5%), more likely to be male (64.4% vs 48.4%), and more likely to have an ECOG performance status <2 (73.0% vs 48.4%) and poor cytogenic risk (30.1% vs 16.1%). Among patients who received first-line systemic therapy who had AML-related mutation(s), *NPM1* (n = 11; 22.4%), MLL^*PTD*^ (n = 11; 22.4%), *CEBPA* (n = 8; 16.3%), *TET2* (n = 8; 16.3%), and *FLT3*^ITD^ (n = 7; 14.3%) were most frequently identified.

Among the 194 patients in this Korean subanalysis, 163 (84.0%) received first-line systemic therapy and 31 (16.0%) received BSC. In the first-line systemic therapy group, 10 had ongoing treatment, 152 discontinued treatment, and the status of one patient was unknown (S1 Fig). There were 145 (89.0%) patients who received HMA monotherapy (azacytidine, n = 5 [3.1%]; decitabine, n = 140 [85.9%]), five (3.1%) who received LDAC monotherapy, and 13 (8.0%) who received HMA and/or LDAC in combination with other systemic therapies (S2 Fig).

### Primary endpoint

Median (95% CI) OS was 7.83 (6.30–9.27) months in patients who received systemic therapy (HMAs, 8.07 [6.27–9.50] months; LDAC and other systemic therapies, 7.57 [3.90–9.80] months), and 4.50 (2.93–11.83) months in those who received BSC (Table 2 and Fig 1). Thirty-seven patients had missing OS data.

**Table 2.**
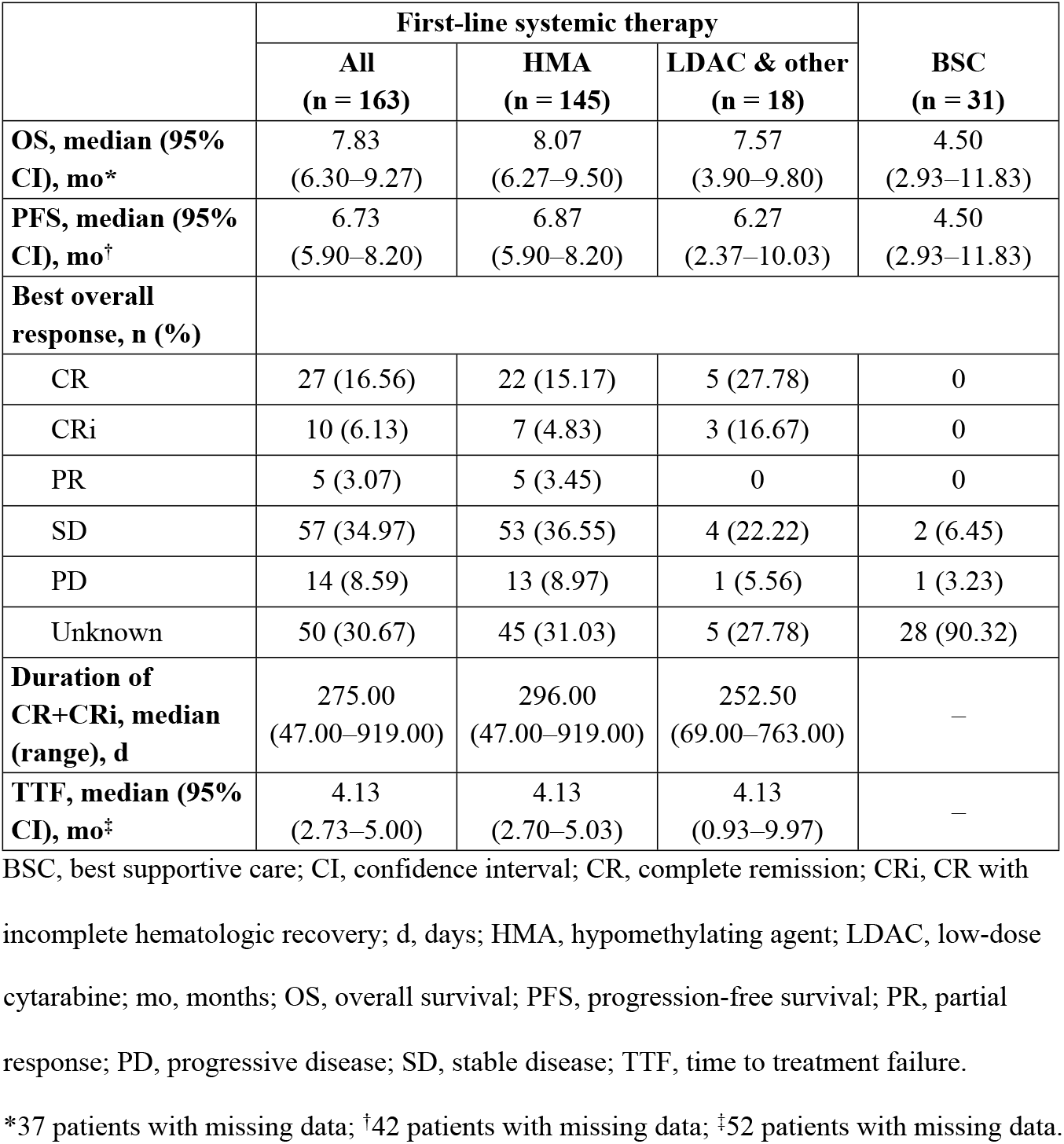
Median OS, PFS, TTF, response rate, and duration of response for patients who received first-line systemic therapy or BSC.

|  | First-line systemic therapy |  |  | BSC<br>(n = 31) |
| --- | --- | --- | --- | --- |
|  | All<br>(n = 163) | HMA<br>(n = 145) | LDAC & other<br>(n = 18) |  |
| **OS, median (95% CI), mo\*** | 7.83<br>(6.30–9.27) | 8.07<br>(6.27–9.50) | 7.57<br>(3.90–9.80) | 4.50<br>(2.93–11.83) |
| **PFS, median (95% CI), mo<sup>†</sup>** | 6.73<br>(5.90–8.20) | 6.87<br>(5.90–8.20) | 6.27<br>(2.37–10.03) | 4.50<br>(2.93–11.83) |
| **Best overall response, n (%)** |  |  |  |  |
| CR | 27 (16.56) | 22 (15.17) | 5 (27.78) | 0 |
| CRi | 10 (6.13) | 7 (4.83) | 3 (16.67) | 0 |
| PR | 5 (3.07) | 5 (3.45) | 0 | 0 |
| SD | 57 (34.97) | 53 (36.55) | 4 (22.22) | 2 (6.45) |
| PD | 14 (8.59) | 13 (8.97) | 1 (5.56) | 1 (3.23) |
| Unknown | 50 (30.67) | 45 (31.03) | 5 (27.78) | 28 (90.32) |
| **Duration of CR+CRi, median (range), d** | 275.00<br>(47.00–919.00) | 296.00<br>(47.00–919.00) | 252.50<br>(69.00–763.00) | – |
| **TTF, median (95% CI), mo<sup>‡</sup>** | 4.13<br>(2.73–5.00) | 4.13<br>(2.70–5.03) | 4.13<br>(0.93–9.97) | – |

BSC, best supportive care; CI, confidence interval; CR, complete remission; CRi, CR with incomplete hematologic recovery; d, days; HMA, hypomethylating agent; LDAC, low-dose
cytarabine; mo, months; OS, overall survival; PFS, progression-free survival; PR, partial response; PD, progressive disease; SD, stable disease; TTF, time to treatment failure. \*37 patients with missing data; †42 patients with missing data; ‡52 patients with missing data.

**Fig 1.**
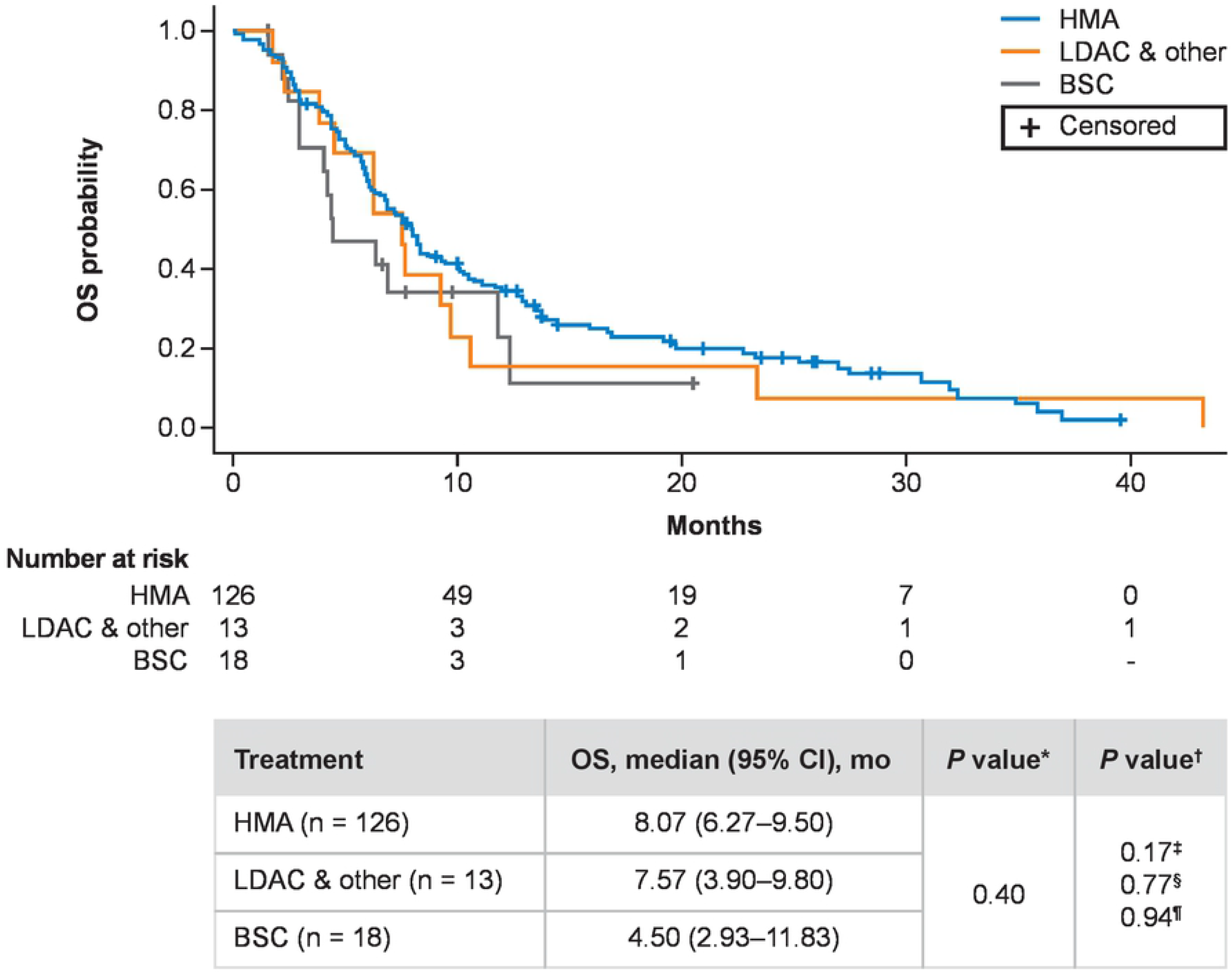
Kaplan-Meier analysis of OS in patients who received HMA, LDAC and other systemic therapies, or BSC. BSC, best supportive care; CI, confidence interval; HMA, hypomethylating agent; LDAC, low-dose cytarabine; mo, months; OS, overall survival. ^*^Log-rank test by comparing between three groups; ^†^Log-rank test by comparing between every two groups; ^‡^HMA vs BSC; ^§^LDAC and other systemic therapies vs BSC; ^¶^HMA vs LDAC and other systemic therapies. Patients with missing data across all groups, n = 37.

Subgroup analyses showed that median OS was significantly different (all *P* <0.005) between patients without (8.20 months) versus with (4.73 months) secondary AML, patients with an ECOG performance status of 0 or 1 (8.30 months) versus ≥2 (4.43 months), patients with favorable (10.67 months) versus intermediate (6.13 months) and poor (6.32 months) cytogenic risk, and patients with CCI of 0 (8.30 months) versus ≥1 (5.73 months; S4 Table).

Using Cox regression analyses, we identified several prognostic factors for OS, including presence of secondary AML (hazard ratio [95% CI], 1.67 [1.13–2.45]; *P* = 0.0094), ECOG performance status ≥2 (2.41 [1.51–3.83]; *P* = 0.0002), intermediate (1.77 [1.10–2.84]; *P* = 0.0182) or poor (2.10 [1.36–3.24]; *P* = 0.0008) cytogenetic risk, and CCI ≥1 (2.26 [1.43–3.58]; *P* = 0.0005; Table 3).

**Table 3.** Prognostic factors that affect OS.

| Category | HR (95% CI) | P value |
| --- | --- | --- |
| **Sex** |  |  |
| Male vs female | 0.98 (0.68–1.41) | 0.9192 |
| **Age** |  |  |
| >75 vs ≤75 years | 0.98 (0.67–1.45) | 0.9279 |
| **Secondary AML** |  |  |
| Yes vs no | 1.67 (1.13–2.45) | 0.0094* |
| Unknown vs no | 2.04 (0.26–16.12) | 0.5006 |
| **ECOG performance status** |  |  |
| ≥2 vs <2 | 2.41 (1.51–3.83) | 0.0002* |
| **Cytogenetic risk<sup>†</sup>** |  |  |
| Intermediate vs favorable | 1.77 (1.10–2.84) | 0.0182* |
| Poor vs favorable | 2.10 (1.36–3.24) | 0.0008* |
| Unknown vs favorable | 2.91 (1.21–6.96) | 0.0167* |
| **Charlson comorbidity index** |  |  |
| ≥1 vs 0 | 2.26 (1.43–3.58) | 0.0005* |

AML, acute myeloid leukemia; CI, confidence interval; ECOG, Eastern Cooperative
Oncology Group; HR, hazard ratio; OS, overall survival.
\*Statistically significant; <sup>†</sup>Risk stratification according to the cytogenetic risk classification
described in S3 Table.

### Secondary endpoints

Median (95% CI) PFS was 6.73 (5.90–8.20) months for patients who received systemic therapy (HMAs, 6.87 [5.90–8.20] months; LDAC and other systemic therapies, 6.27 [2.37–10.03] months), and 4.50 (2.93–11.83) months for patients who received BSC (Table 2 and S3 Fig). Median (95% CI) TTF was 4.13 (2.73–5.00) months for patients who received systemic therapy (HMAs: 4.13 [2.70–5.03] months; LDAC and other systemic therapies: 4.13 [0.93–9.97] months; Table 2). The number of patients with missing data for PFS and TTF was 42 and 52, respectively. Among the 163 patients who received systemic therapy, 37 (22.7%) achieved CR or CRi, with a median (95% CI) DoR of 275.00 (47.00–919.00) days (Table 2). CR or CRi was achieved in 20.0% of patients who received HMAs and 44.4% of patients who received LDAC and other systemic therapies, with a corresponding median (95% CI) DoR of 296.00 (47.00–919.00) and 252.50 (69.00–763.00) days, respectively (Table 2).

Subgroup analysis showed that median PFS was significantly different (all *P* <0.05) between patients without (7.37 months) versus with (4.68 months) secondary AML, patients with ECOG performance status 0 or 1 (7.23 months) versus ≥2 (4.20 months), patients with favorable (8.37 months) versus intermediate (5.77 months) and poor (6.23 months) cytogenic risk, and patients with CCI of 0 (7.27 months) versus ≥1 (5.73 months; S5 Table).

Using Cox regression analyses, we identified several factors associated with PFS, including presence of secondary AML (hazard ratio [95% CI], 1.58 [1.08–2.33]; *P* = 0.0190), ECOG performance status ≥2 (2.25 [1.40–3.62]; *P* = 0.0008), poor cytogenetic risk (1.96 [1.27–3.04]; *P* = 0.0026), and CCI ≥1 (2.01 [1.28–3.16]; *P* = 0.0025; S6 Table).

## Discussion

In the overall CURRENT study population, HMAs were associated with longer median OS, PFS, and TTF, compared with other systemic therapies or BSC [7]. This subanalysis revealed similar survival outcomes among the study’s Korean subpopulation. We also found that several patient demographic and genetic factors were associated with OS and PFS.

Survival outcomes among all patients in this Korean subanalysis were poor. Median OS was higher in patients who received systemic therapy (7.83 months) compared with those who received BSC (4.50 months), although this was not statistically significant. Notably, median OS was highest in patients who received HMAs (8.07 months). Survival outcomes in patients receiving HMAs were largely consistent with previous reports in clinical trials [17-19] and real-world studies [20] (median OS, 6.6–10.4 months). In line with previous studies and the overall CURRENT study [16], this subanalysis highlights the preference for HMAs in patients who are ineligible to receive ICT, which was not surprising given the favorable survival outcomes associated with HMAs compared with other available therapies. Median OS for the HMA cohort in this subanalysis closely mirrored that of a systematic review and meta-analysis of the efficacy and safety of decitabine in the treatment of elderly patients with AML (n = 718; median [95% CI] OS, 8.09 [5.77–10.41] months) [21]. Notably, 85.9% of patients in the present subanalysis received decitabine as first-line systemic therapy. In contrast to our results, a US study reported a median (95% CI) OS of 4.30 (3.20–5.80) months in patients treated with HMAs [22]. Furthermore, median OS in the LDAC and BSC cohorts were slightly longer than reported previously [18]. These differences may be explained by the present population being more representative of real-world clinical practice and comprising only Korean patients. When evaluating OS in patient subgroups, we found that those diagnosed with versus without secondary AML, with an ECOG performance status ≥2 versus 0 or 1, with poor or intermediate versus favorable cytogenetic risk, or with CCI ≥1 versus 0 had a shorter median OS. Similar observations have been reported in previous studies [14, 23-25].

Median PFS in this subanalysis was higher in patients who received systemic therapy (6.73 months) compared with patients who received BSC (4.50 months), although this was not statistically significant. Notably, median PFS was highest in patients who received HMAs (6.87 months), which is consistent with the global CURRENT study [16]. Evaluation of PFS according to patient subgroups revealed that patients diagnosed with versus without secondary AML, with an ECOG performance status ≥2 versus 0 or 1, with poor or intermediate versus favorable cytogenetic risk, or with CCI ≥1 versus 0 had shorter median PFS. These results are consistent with previous studies in which poor ECOG performance status and comorbidity index scores were associated with shorter median PFS [23].

Median TTF was comparable between all patients receiving first-line systemic therapies, which is in contrast to the overall CURRENT study in which longer median TTF was reported in patients who received HMAs [16]. CR and CRi rates were lower in patients who received HMAs compared with LDAC and other systemic therapies, which is consistent with results from the CURRENT study [16]. On the other hand, median duration of CR and CRi were higher in patients who received HMAs compared with other systemic therapies, which was not observed in the main study [16].

Baseline characteristics of Korean patients in this subanalysis were generally consistent with the global CURRENT study [16]. The vast majority of patients reported comorbidities, and patients who received HMAs were more likely to report ECOG performance status <2 with favorable or intermediate cytogenic risk, compared with patients who received LDAC and other systemic therapies, or BSC. The mutation rate in this subanalysis among patients who received systemic therapies was 34.0%; the most frequently occurring mutations reported here and in the CURRENT study [16] were *NPM1* and *FLT3*^ITD^, confirming findings from a previous report [26]. In addition, we found that there was a significant difference between median age, proportion of male patients, and the proportion of patients aged >75 years for the HMA, LDAC and other systemic therapies, and BSC groups. Fewer patients in the systemic therapies groups versus the BSC group were >75 years of age, indicating that patients in this subanalysis who received systemic therapies may have had a better prognosis [27], although age was not found to be a significant prognostic factor for survival in this Korean subanalysis.

Factors associated with poorer OS and PFS included secondary AML, ECOG performance status ≥2, intermediate or poor cytogenetic risk, and CCI ≥1. This is consistent with a multicenter trial in which better performance status, non-adverse cytogenetics, and lower CCI scores were associated with better survival outcomes in patients with AML who were ineligible for ICT and received decitabine as first-line treatment [28]. Better performance status was similarly found to be prognostic for survival in elderly Korean patients with AML [29]. This may have influenced the outcomes of patients in our study, in which 75% of patients in the HMA group had ECOG performance status <2 compared with just 55.6% and 48.4% in the LDAC and other systemic therapies and BSC groups, respectively. In contrast to our results, a study of 248 elderly patients on low-intensity therapy did not find an association between survival and ECOG performance status or cytogenetic risk, but identified response to the first induction cycle and lactate dehydrogenase levels as prognostic parameters [27], neither of which were examined in our study. With regard to treatment with HMAs, patients with DNA methylation-related mutations have improved OS, and *TET2* mutation has been recognized as an independent prognostic factor for PFS [30]. In this subanalysis, *TET2* mutation was identified in 18.2% of patients in the HMA cohort, whereas none of the patients in the other treatment groups had this mutation. Overall, the prognostic parameters associated with median OS and PFS in our study were consistent with those reported in patients who received ICT [19, 31-34].

Finally, we have shown that more patients who were ineligible for ICT received HMAs compared with LDAC and BSC, which is consistent with the CURRENT study [16]. Regardless, survival was poor among all patients. Studies investigating outcomes in patients who received HMA compared with ICT have found that HMA was more frequently used in older patients, despite better outcomes with ICT, even in those with comorbidities [20, 22]. Conversely, two recent analyses of elderly patients (≥65 years) with AML in Korea noted that despite lower response rates in patients who received HMAs compared with those who received ICT, survival outcomes were comparable [35, 36]. Other studies involving elderly patients with AML have also reported comparable or better survival outcomes for those who received HMAs compared with those who received ICT or palliative care [37]. Notably, there were patients in this subanalysis who received only palliative BSC despite the availability of first-line systemic therapies. Given that baseline characteristics, except for age, were largely consistent between the first-line systemic therapy and BSC groups, it may be that BSC is considered for elderly patients because age is regarded as a critical factor when making treatment decisions. There remains a significant unmet need for higher efficacy treatments for patients who are ineligible for ICT owing to advanced age. Although targeted treatments have been associated with a moderate improvement in outcomes for patients unfit for ICT [38-43], prognosis remains poor and there is a lack of consensus regarding optimal treatment for these patients.

Several limitations should be considered when interpreting the results of this study. As with all real-world retrospective studies, the CURRENT study was uncontrolled and nonrandomized. Missing data may limit interpretation; missing molecular and cytogenetic data may limit assessment of their effect on outcomes, and missing response rate data for >30% of patients who received systemic therapies may limit the generalizability of these findings. There are many systemic therapies included in the “other systemic therapy” group of this study, which may limit interpretation of the clinical outcomes of patients who received each of these therapies. Intra- and inter-site variability may exist, but to reduce variations and the need for corrections in the data collected, we optimized and ensured the clarity of the electronic CRF, and provided all study sites with adequate training.

## Conclusion

Overall, this subanalysis of the real-world CURRENT study provided several insights into the clinical management of Korean patients with AML who are ineligible for ICT. The clinical outcomes for this Korean subgroup are poor, with a median OS <10 months in patients who received systemic therapy and <5 months in patients who received BSC. The majority of Korean patients with AML who are unfit for ICT receive HMAs, which are associated with numerically longer median OS and PFS relative to other systemic therapies and BSC. Factors such as secondary AML, ECOG performance status, cytogenetic risk, and CCI may be prognostic for survival. Given the rising incidence of AML due to the aging population, there is a substantial unmet need for novel therapies and combination regimens to improve clinical outcomes in this patient population.

## Data Availability

All relevant data are within the manuscript. The data presented in this study are available from AbbVie Inc., North Chicago, IL, USA (contact via). Authors share the “minimal data set” for their submission. The minimal data set to consist of the data required to replicate all study findings reported in the article, as well as related metadata and methods. Additionally, authors comply with field-specific standards for preparation, recording, and deposition of data when applicable.

## Acknowledgments

Editorial assistance was provided by Liting Hang BSc (Hons), PhD, and Alice Carruthers BSc (Hons), PhD, (Nucleus Global Shanghai, Shanghai, China) and statistical analysis was provided by Medi Help Line, both funded by AbbVie Korea. We would like to acknowledge the contributions of our colleague, Maria Belen Guijarro Garbayo in memoriam, formerly of AbbVie, for her contributions to the study concept and design, and contributions to drafts of this publication.

## Supporting information

**S1 Table. Baseline comorbidities**. Each patient can have multiple comorbidities. BSC, best supportive care; HMA, hypomethylating agent; LDAC, low-dose cytarabine. *Fisher exact test; ^†^Marked as ‘Other’ in case report form (disease not specified). *P* value indicates statistical difference in a three-way comparison between BSC, HMA, and LDAC and other systemic therapies.

**S2 Table. Baseline molecular profiling and cytogenetic risk**. Percentages may total more than 100% because multiselection was allowed. BSC, best supportive care; HMA, hypomethylating agent; LDAC, low-dose cytarabine; NGS, next-generation sequencing.

^*^Chi-squared test; ^**^Kruskal-Wallis test; ^†^Percentages were calculated for each respective treatment group using the number of patients with known mutation status from NGS or targeted mutation testing; ^‡^Risk stratification according to the cytogenetic risk classification described in S3 Table. *P* value indicates statistical difference in a three-way comparison between BSC, HMA, and LDAC and other systemic therapies.

**S3 Table. Cytogenetic risk classification**.

**S4 Table. Kaplan-Meier estimate for median OS by baseline clinical characteristics**. AML, acute myeloid leukemia; AML-MRC, AML with myelodysplasia-related changes; BSC, best supportive care; CI, confidence interval; ECOG, Eastern Cooperative Oncology Group; mo, months; NA, not applicable; NR, not reached; OS, overall survival. *Statistically significant; ^†^Risk stratification according to the cytogenetic risk classification described in S3 Table. The lower limit of the 95% CI is a closed interval (indicated by a square bracket) whereas the upper limit of the 95% CI is an open interval (indicated by a round bracket).

**S5 Table. Kaplan-Meier estimate for median PFS by baseline clinical characteristics**. AML, acute myeloid leukemia; AML-MRC, AML with myelodysplasia-related changes; BSC, best supportive care; CI, confidence interval; ECOG, Eastern Cooperative Oncology Group; mo, months; NA, not applicable; NR, not reached; PFS, progression-free survival.

*Statistically significant; ^†^Risk stratification according to the cytogenetic risk classification described in S3 Table. The lower limit of the 95% CI is a closed interval (indicated by a square bracket) whereas the upper limit of the 95% CI is an open interval (indicated by a round bracket).

**S6 Table. Prognostic factors that affect PFS**. AML, acute myeloid leukemia; CI, confidence interval; ECOG, Eastern Cooperative Oncology Group; HR, hazard ratio, PFS, progression-free survival. *Statistically significant.

**S1 Fig. Patient disposition**. ^*^Overlapping count.

**S2 Fig. Overview of patients receiving first-line systemic therapies and BSC**. BSC, best supportive care; CA ± G, cytarabine and aclarubicin ± granulocyte colony-stimulating factor combination regimen; LDAC, low-dose cytarabine.

**S3 Fig. Kaplan–Meier analysis of PFS in patients who received HMA, LDAC and other systemic therapies, and BSC**. BSC, best supportive care; CI, confidence interval; HMA, hypomethylating agent; LDAC, low-dose cytarabine; mo, months; PFS, progression-free survival. ^*^Log-rank test by comparing between three groups; ^†^Log-rank test by comparing between every two groups; ^‡^HMA vs BSC; ^§^LDAC & other vs BSC; ^¶^HMA vs LDAC & other. Patients with missing data across all groups, n = 42.

## References

1. Saultz JN, Garzon R. Acute myeloid leukemia: A concise review. J Clin Med. 2016;5(3):33.

2. Pollyea DA, Bixby D, Perl A, Bhatt VR, Altman JK, Appelbaum FR, et al. NCCN guidelines insights: Acute myeloid leukemia, Version 2. 2021. J Natl Compr Canc Netw. 2021;19(1):16–27.

3. Cancer stat facts: Leukemia - acute myeloid leukemia. National Cancer Institute Surveillance, Epidemiology and End Results Program. 2021. Available from: https://seer.cancer.gov/statfacts/html/amyl.html.

4. Yi M, Li A, Zhou L, Chu Q, Song Y, Wu K. The global burden and attributable risk factor analysis of acute myeloid leukemia in 195 countries and territories from 1990 to 2017: Estimates based on the global burden of disease study 2017. J Hematol Oncol. 2020;13(1):72.

5. Park E-H, Lee H, Won Y-J, Ju HY, Oh C-M, Ingabire C, et al. Nationwide statistical analysis of myeloid malignancies in Korea: Incidence and survival rate from 1999 to 2012. Blood Res. 2015;50(4):204–17.

6. Thein MS, Ershler WB, Jemal A, Yates JW, Baer MR. Outcome of older patients with acute myeloid leukemia: an analysis of SEER data over 3 decades. Cancer. 2013;119(15):2720–7.

7. Deschler B, de Witte T, Mertelsmann R, Lübbert M. Treatment decision-making for older patients with high-risk myelodysplastic syndrome or acute myeloid leukemia: problems and approaches. Haematologica. 2006;91(11):1513–22.

8. Ferrara F, Barosi G, Venditti A, Angelucci E, Gobbi M, Pane F, et al. Consensus-based definition of unfitness to intensive and non-intensive chemotherapy in acute myeloid leukemia: a project of SIE, SIES and GITMO group on a new tool for therapy decision making. Leukemia. 2013;27(5):997–9.

9. Döhner H, Estey E, Grimwade D, Amadori S, Appelbaum FR, Büchner T, et al. Diagnosis and management of AML in adults: 2017 ELN recommendations from an international expert panel. Blood. 2017;129(4):424–47.

10. Heuser M, Ofran Y, Boissel N, Brunet Mauri S, Craddock C, Janssen J, et al. Acute myeloid leukaemia in adult patients: ESMO clinical practice guidelines for diagnosis, treatment and follow-up. Ann Oncol. 2020;31(6):697–712.

11. Palmieri R, Paterno G, De Bellis E, Mercante L, Buzzatti E, Esposito F, et al. Therapeutic choice in older patients with acute myeloid leukemia: A matter of fitness. Cancers (Basel). 2020;12(1):120.

12. Büchner T, Berdel WE, Haferlach C, Haferlach T, Schnittger S, Müller-Tidow C, et al. Age-related risk profile and chemotherapy dose response in acute myeloid leukemia: a study by the German Acute Myeloid Leukemia Cooperative Group. J Clin Oncol. 2009;27(1):61–9.

13. Wahlin A, Markevärn B, Golovleva I, Nilsson M. Prognostic significance of risk group stratification in elderly patients with acute myeloid leukaemia. Br J Haematol. 2001;115(1):25–33.

14. Wheatley K, Brookes CL, Howman AJ, Goldstone AH, Milligan DW, Prentice AG, et al. Prognostic factor analysis of the survival of elderly patients with AML in the MRC AML11 and LRF AML14 trials. Br J Haematol. 2009;145(5):598–605.

15. Krok-Schoen JL, Fisher JL, Stephens JA, Mims A, Ayyappan S, Woyach JA, et al. Incidence and survival of hematological cancers among adults ages ≥75 years. Cancer Med. 2018;7(7):3425–33.

16. Miyamoto T, Sanford D, Tomuleasa C, Hsiao HH, Olivera LJE, Enjeti AK, et al. Real-world treatment patterns and clinical outcomes in patients with AML unfit for first-line intensive chemotherapy. Leuk Lymphoma. 2022;63(4):928–38.

17. Dombret H, Seymour JF, Butrym A, Wierzbowska A, Selleslag D, Jang JH, et al. International phase 3 study of azacitidine vs conventional care regimens in older patients with newly diagnosed AML with >30% blasts. Blood. 2015;126(3):291–9.

18. Seymour JF, Döhner H, Butrym A, Wierzbowska A, Selleslag D, Jang JH, et al. Azacitidine improves clinical outcomes in older patients with acute myeloid leukaemia with myelodysplasia-related changes compared with conventional care regimens. BMC Cancer. 2017;17(1):852.

19. Kantarjian HM, Thomas XG, Dmoszynska A, Wierzbowska A, Mazur G, Mayer J, et al. Multicenter, randomized, open-label, phase III trial of decitabine versus patient choice, with physician advice, of either supportive care or low-dose cytarabine for the treatment of older patients with newly diagnosed acute myeloid leukemia. J Clin Oncol. 2012;30(21):2670–7.

20. Medeiros BC, Satram-Hoang S, Hurst D, Hoang KQ, Momin F, Reyes C. Big data analysis of treatment patterns and outcomes among elderly acute myeloid leukemia patients in the United States. Ann Hematol. 2015;94(7):1127–38.

21. He P-F, Zhou J-D, Yao D-M, Ma J-C, Wen X-M, Zhang Z-H, et al. Efficacy and safety of decitabine in treatment of elderly patients with acute myeloid leukemia: A systematic review and meta-analysis. Oncotarget. 2017;8(25):41498–507.

22. Bell JA, Galaznik A, Farrelly E, Blazer M, Murty S, Ogbonnaya A, et al. A retrospective study evaluating treatment patterns and survival outcomes in elderly patients with acute myeloid leukemia treated in the United States with either 7+3 or a hypomethylating agent. Leuk Res. 2019;78:45–51.

23. Papageorgiou SG, Kotsianidis I, Bouchla A, Symeonidis A, Galanopoulos A, Viniou N-A, et al. Serum ferritin and ECOG performance status predict the response and improve the prognostic value of IPSS or IPSS-R in patients with high-risk myelodysplastic syndromes and oligoblastic acute myeloid leukemia treated with 5- azacytidine: a retrospective analysis of the Hellenic national registry of myelodysplastic and hypoplastic syndromes. Ther Adv Hematol. 2020;11:2040620720966121.

24. Dhakal P, Shostrom V, Al-Kadhimi ZS, Maness LJ, Gundabolu K, Bhatt VR. Usefulness of Charlson comorbidity index to predict early mortality and overall survival in older patients with acute myeloid leukemia. Clin Lymphoma Myeloma Leuk. 2020;20(12):804–12.e8.

25. Gbadamosi B, Ezekwudo D, Bastola S, Jaiyesimi I. Predictive and prognostic markers in adults with acute myeloid leukemia: A single-institution experience. Clin Lymphoma Myeloma Leuk. 2018;18(7):e287–e94.

26. Papaemmanuil E, Gerstung M, Bullinger L, Gaidzik VI, Paschka P, Roberts ND, et al. Genomic classification and prognosis in acute myeloid leukemia. The New England journal of medicine. 2016;374(23):2209–21.

27. Chen Y, Yang T, Zheng X, Yang X, Zheng Z, Zheng J, et al. The outcome and prognostic factors of 248 elderly patients with acute myeloid leukemia treated with standard-dose or low-intensity induction therapy. Medicine (Baltimore). 2016;95(30):e4182.

28. Lübbert M, Rüter BH, Claus R, Schmoor C, Schmid M, Germing U, et al. A multicenter phase II trial of decitabine as first-line treatment for older patients with acute myeloid leukemia judged unfit for induction chemotherapy. Haematologica. 2012;97(3):393–401.

29. Yi HG, Lee MH, Kim CS, Hong J, Park J, Lee JH, et al. Clinical characteristics and treatment outcome of acute myeloid leukemia in elderly patients in Korea: A retrospective analysis. Blood Res. 2014;49(2):95–9.

30. Wang RQ, Chen CJ, Jing Y, Qin JY, Li Y, Chen GF, et al. Characteristics and prognostic significance of genetic mutations in acute myeloid leukemia based on a targeted next-generation sequencing technique. Cancer Med. 2020;9(22):8457–67.

31. Roboz GJ, Wei AH, Ravandi F, Pocock C, Montesinos P, Dombret H, et al. Prognostic factors of overall (OS) and relapse-free survival (RFS) for patients with acute myeloid leukemia (AML) in remission after intensive chemotherapy (IC): Multivariate analyses from the QUAZAR AML-001 trial of oral azacitidine (Oral- AZA). J Clin Oncol. 2021;39(15_Suppl):7014.

32. Kim DS, Kang KW, Yu ES, Kim HJ, Kim JS, Lee SR, et al. Selection of elderly acute myeloid leukemia patients for intensive chemotherapy: effectiveness of intensive chemotherapy and subgroup analysis. Acta Haematol. 2015;133(3):300–9.

33. Ma TT, Lin XJ, Cheng WY, Xue Q, Wang SY, Liu FJ, et al. Development and validation of a prognostic model for adult patients with acute myeloid leukaemia. EBioMedicine. 2020;62:103126.

34. Suvajdžić N, Cvetković Z, Dorđević V, Kraguljac-Kurtović N, Stanisavljević D, Bogdanović A, et al. Prognostic factors for therapy-related acute myeloid leukaemia (t-AML)--a single centre experience. Biomed Pharmacother. 2012;66(4):285–92.

35. Oh SB, Park SW, Chung JS, Lee WS, Lee HS, Cho SH, et al. Therapeutic decision- making in elderly patients with acute myeloid leukemia: conventional intensive chemotherapy versus hypomethylating agent therapy. Ann Hematol. 2017;96(11):1801–9.

36. Choi EJ, Lee JH, Park HS, Lee JH, Seol M, Lee YS, et al. Decitabine versus intensive chemotherapy for elderly patients with newly diagnosed acute myeloid leukemia. Clin Lymphoma Myeloma Leuk. 2019;19(5):290–9.e3.

37. Österroos A, Eriksson A, Antunovic P, Cammenga J, Deneberg S, Lazarevic V, et al. Real-world data on treatment patterns and outcomes of hypomethylating therapy in patients with newly diagnosed acute myeloid leukaemia aged ≥ 60 years. Br J Haematol. 2020;189(1):e13–e6.

38. Amadori S, Suciu S, Selleslag D, Aversa F, Gaidano G, Musso M, et al. Gemtuzumab ozogamicin versus best supportive care in older patients with newly diagnosed acute myeloid leukemia unsuitable for intensive chemotherapy: Results of the randomized phase III EORTC-GIMEMA AML-19 trial. J Clin Oncol. 2016;34(9):972–9.

39. Cortes JE, Heidel FH, Hellmann A, Fiedler W, Smith BD, Robak T, et al. Randomized comparison of low dose cytarabine with or without glasdegib in patients with newly diagnosed acute myeloid leukemia or high-risk myelodysplastic syndrome. Leukemia. 2019;33(2):379–89.

40. DiNardo CD, Pratz K, Pullarkat V, Jonas BA, Arellano M, Becker PS, et al. Venetoclax combined with decitabine or azacitidine in treatment-naive, elderly patients with acute myeloid leukemia. Blood. 2019;133(1):7–17.

41. Lambert J, Pautas C, Terré C, Raffoux E, Turlure P, Caillot D, et al. Gemtuzumab ozogamicin for de novo acute myeloid leukemia: final efficacy and safety updates from the open-label, phase III ALFA-0701 trial. Haematologica. 2019;104(1):113–9.

42. Ohanian M, Garcia-Manero G, Levis M, Jabbour E, Daver N, Borthakur G, et al. Sorafenib combined with 5-azacytidine in older patients with untreated FLT3-ITD mutated acute myeloid leukemia. Am J Hematol. 2018;93(9):1136–41.

43. Pollyea DA, Tallman MS, de Botton S, Kantarjian HM, Collins R, Stein AS, et al. Enasidenib, an inhibitor of mutant IDH2 proteins, induces durable remissions in older patients with newly diagnosed acute myeloid leukemia. Leukemia. 2019;33(11):2575–84.

